# Syncope at SARS-CoV-2 onset due to impaired baroreflex response

**DOI:** 10.1101/2020.05.29.20114751

**Authors:** Ciro Canetta, Silvia Accordino, Elisabetta Buscarini, Gianpaolo Benelli, Giuseppe La Piana, Alessandro Scartabellati, Giovanni Viganò, Roberto Assandri, Alberto Astengo, Chiara Benzoni, Gianfranco Gaudiano, Daniele Cazzato, Davide Sebastiano Rossi, Susanna Usai, Irene Tramacere, Giuseppe Lauria

**Author notes:** Corresponding Author: Giuseppe Lauria, MD, Department of Biomedical and Clinical Sciences “Luigi Sacco”, University of Milan, Department of Clinical Neuroscience, IRCCS Foundation “Carlo Besta” Neurological Institute Milan, Italy.

## Abstract

We describe clinical and laboratory findings in 35 consecutive patients tested positive for SARS-CoV-2 by reverse transcriptase-polymerase chain reaction on nasopharyngeal swab that presented one or multiple syncopal events at disease onset. Neurological examination and electrocardiographic findings were normal. Chest computed tomography showed findings consistent with interstitial pneumonia. Arterial blood gas analysis showed low pO2, pCO2, and P/F ratio indicating hypocapnic hypoxemia, while patients did not show the expected compensatory heart rate increase. Such mechanism could have led to syncope. We speculate that SARS-CoV-2 could have caused angiotensin-converting enzyme-2 (ACE2) receptor internalization in the nucleus of the solitary tract (NTS), thus altering the baroreflex response and inhibiting the compensatory tachycardia during acute hypocapnic hypoxemia.

---

Since the beginning of the severe acute respiratory syndrome coronavirus 2 (SARS-CoV-2) pandemic in Italy on February 19^th^, 2020, the Lombardy Region in Northern Italy has been one the most affected areas in Europe. The public hospital in the town of Crema was one of the first to face the exponential influx of patients. Thanks to the immediate adoption of available local procedures to cope with the hospitalization of patients with a potential viral spread, based on 2009 SARS and H1N1 pandemic strategic plan revised on December 2014 after Ebola outbreak, the Emergency Department could set up a standardized triage for any individual either reporting or presenting with fever, cough, or dyspnea, or having had contact with potentially Covid-19 carriers.

## METHODS

Since February 21^th^, all consecutive suspected patients admitted to the hospital underwent a procedure including body temperature and pulse oximetry (SO2) recording, hematological screening, chest X-ray and/or computed tomography (CT) scan, and nasopharyngeal swab. Swabs were stored at +4°C and immediately shipped to one of the laboratory of virology accredited by the Lombardy Region to perform diagnostic SARS-COV-2 real-time polymerase chain reaction (RT-PCR) assay. Based on the clinical, laboratory, and radiological findings, patients were discharged to home in quarantine or admitted to the hospital. All necessary patient consent has been obtained and the appropriate institutional forms have been archived.

Recently, some case series have reported syncope as the first manifestation of SARS-CoV-2 infection, even in the absence of other common symptoms such as fever, cough, and dyspnea. Patients had in common comorbidities for heart disease, coronary artery disease, post-permanent pacemaker (PPM) or cardiac loop recorder implantation (Ebrille et al., 2020; Tape et al., 2020). We analyzed 411 consecutive patients tested positive for SARS-CoV-2 by reverse transcriptase-polymerase chain reaction at nasopharyngeal swab among whom nearly 10% reported syncope at the onset of the infection (Benelli et al., 2020). Herewith we describe clinical and laboratory findings in 35 consecutive patients that presented one or multiple (9 patients; 25.7%) syncopal events at disease onset.

Syncope occurred within 3.6±2.7 days from hospital admission. Nine (25.7%) patients reported head trauma and one subdural haemorrhage. Associated onset symptoms were fever >37.5°C in 17 (48.5%) patients, cough in 8 (22.8%), and dyspnoea in 7 (20%). Hypertension (45.7%), dyslipidaemia (17%), renal insufficiency, hypothyroidism, dementia, cancer (8.5%), anaemia and atrial fibrillation (5.7%) were the most common comorbidities. Twenty-one (60%) patients reported intake of >3 drugs, 12 (34.3%) of 4-6 drugs and 2 (5.7%) of >7 drugs. Eleven (31.4%) patients were taking beta-blockers.

The neurological examination was normal in all patients. Chest computed tomography showed single or multiple ground-glass and/or consolidative lung opacities consistent with interstitial pneumonia in all patients. Electrocardiogram showed corrected QT interval at rest ranging 409-511 ms (mean 451±30; median 449.5; interquartile range 433-474), PR interval ranging 104-220 ms (mean 161±35.5; median 153; interquartile range 136.5-195.5), and QRS complex duration ranging 66-134 ms (mean 95±18; median 93; interquartile range 83-105.5), which were nearly within normal values. Initial arterial blood gas analysis showed low pO2, pCO2, and P/F ratio consistent with hypocapnic hypoxemia in most patients (table 1). However, patients did not show compensatory heart rate increase (fig. 1). Such mechanism likely underlay the occurrence of syncope (Yasuma et al., 2000).

**Table 1.**
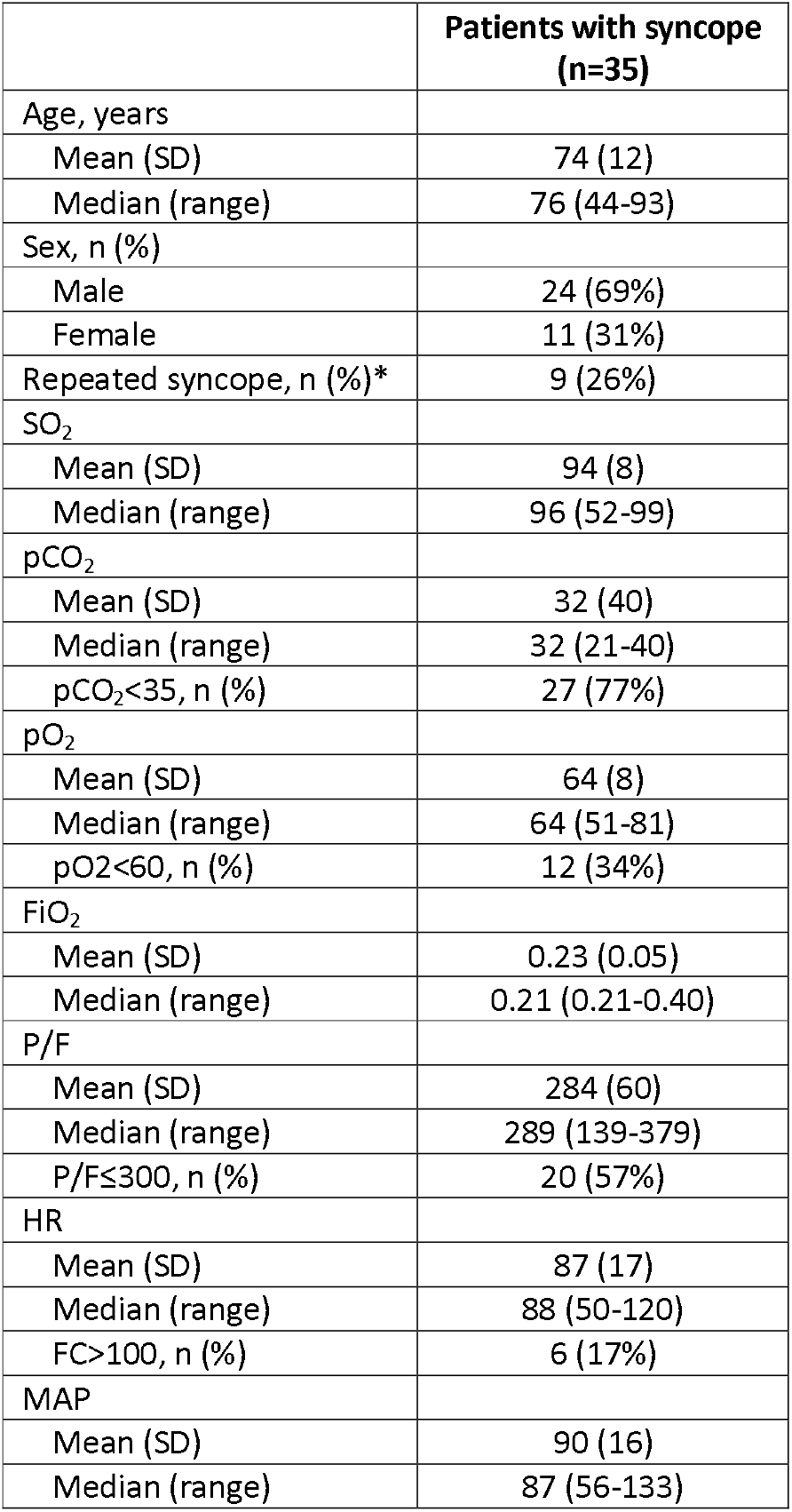
SARS-Cov-2 patients presenting with syncope initial features. SO_2_ is oxygen saturation; pCO2 and pO2 are partial pressure of CO2 and O2; FiO2 is fraction of inspired oxygen; P/F is the ratio of arterial oxygen partial pressure (PaO2) to fractional inspired oxygen (FiO2); HR is heart rate; MAP is mean arterial pressure [diastolic + l/3(systolic-diastolic)].

|  | Patients with syncope<br>(n=35) |
| --- | --- |
| Age, years |  |
| Mean (SD) | 74 (12) |
| Median (range) | 76 (44-93) |
| Sex, n (%) |  |
| Male | 24 (69%) |
| Female | 11 (31%) |
| Repeated syncope, n (%)* | 9 (26%) |
| SO<sub>2</sub> |  |
| Mean (SD) | 94 (8) |
| Median (range) | 96 (52-99) |
| pCO<sub>2</sub> |  |
| Mean (SD) | 32 (40) |
| Median (range) | 32 (21-40) |
| pCO<sub>2</sub><35, n (%) | 27 (77%) |
| pO<sub>2</sub> |  |
| Mean (SD) | 64 (8) |
| Median (range) | 64 (51-81) |
| pO<sub>2</sub><60, n (%) | 12 (34%) |
| FiO<sub>2</sub> |  |
| Mean (SD) | 0.23 (0.05) |
| Median (range) | 0.21 (0.21-0.40) |
| P/F |  |
| Mean (SD) | 284 (60) |
| Median (range) | 289 (139-379) |
| P/F≤300, n (%) | 20 (57%) |
| HR |  |
| Mean (SD) | 87 (17) |
| Median (range) | 88 (50-120) |
| FC>100, n (%) | 6 (17%) |
| MAP |  |
| Mean (SD) | 90 (16) |
| Median (range) | 87 (56-133) |

**Figure 1.**
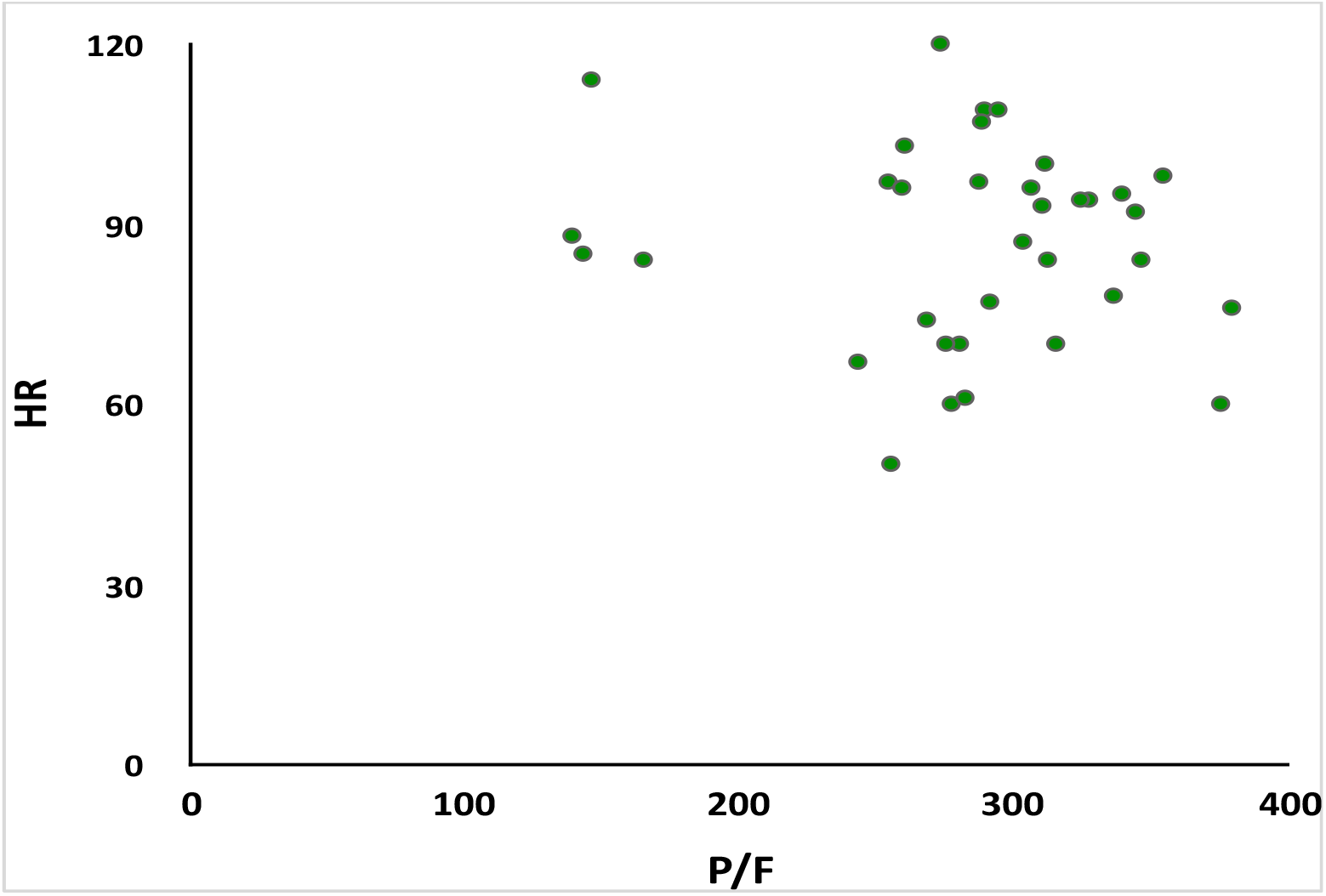
Scatter plot showing no correlation (rho=-0.04, p=0.84) between heart rate (HR) and ratio of (pO2) arterial oxygen partial pressure to (FiO2) fractional inspired oxygen (P/F).

Cardiovascular and respiratory systems strongly interact to ensure adequate oxygen release to tissues and organs, and the balance between sympathetic and parasympathetic inputs predominantly regulates heart rate response. The increased compensatory tachycardia response to hypoxia induces higher cardiac output, while stroke volume remains unchanged.

SARS-CoV-2 viral coat expresses the S protein that contains a receptor-binding region with high affinity for the extracellular domain of angiotensin-converting enzyme-2 (ACE2) receptor. Binding leads to ACE2 internalization. It has been shown that the loss of ACE2 at cell surface could precipitate existing cardiovascular, kidney and brain diseases (South et al., 2020). The baroreceptor reflex control of heart rate is regulated by the brain renin-angiotensin system in the nucleus of the solitary tract (NTS) of the brainstem, where ACE2 is expressed (Doobay et al., 2007). Increased ACE2 activity in the brainstem decreases baroreflex sensitivity and vagal tone, and increases sympathetic output and heart rate (Mukerjee et al., 2019). Conversely, experimental injection of ACE2 antagonist in rat NTS increased sensitivity of the baroreceptor reflex leading to bradycardia response (Diz et al., 2008).

In patients experiencing syncope, SARS-CoV-2 could have caused ACE2 internalization in the NTS, thus altering the baroreflex response and inhibiting the compensatory increase of heart rate during acute.

## Data Availability

See Availability statement

## Author Guarantee Statement

Kindly confirm that your manuscript meets the following criteria by marking an ‘x’ in each appropriate box on the left. Alternatively, print the form, fill it in and scan it for submission with your manuscript files.

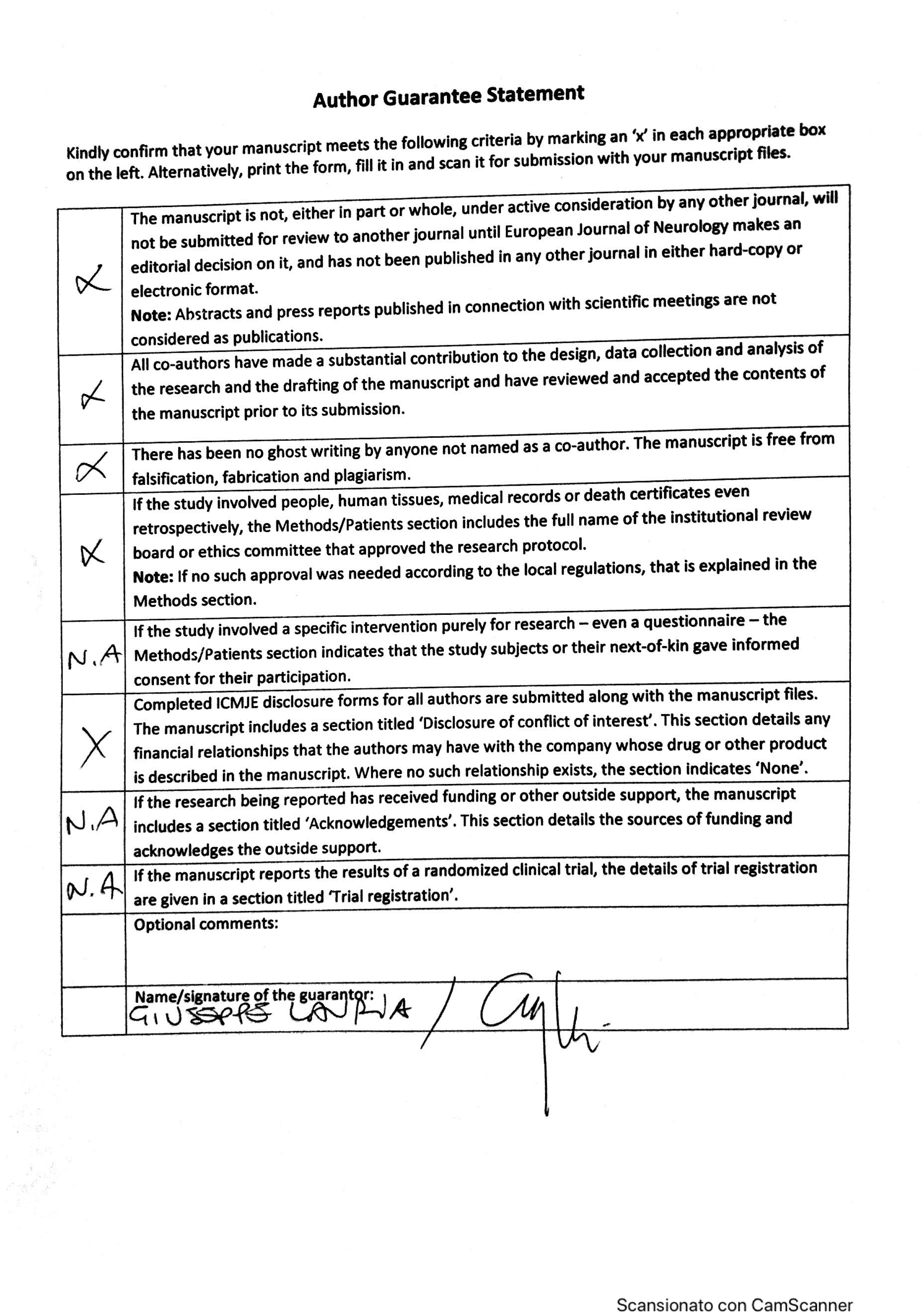

